# Targeted Binding of Nitrogenous Waste Products Using Antibody-Coated Granules: A New Approach for CKD Management

**DOI:** 10.64898/2026.04.28.26351724

**Authors:** Sherif Salah Abdul Aziz, Abdullah Mubarki, Mohamed Sherif Salah

**Author notes:** Corresponding Authors: <u>Dr.</u> Sherif Salah Abdul Aziz.

## Abstract

Chronic kidney disease is a progressive condition characterized by the accumulation of nitrogenous waste products, including urea, creatinine, and uric acid, leading to significant morbidity in advanced stages. Current management strategies, such as dialysis, are effective but associated with substantial clinical and socioeconomic burdens, highlighting the need for alternative approaches to reduce circulating toxins.

In this study, we evaluated a novel formulation of psyllium-based granules functionalized with specific antibody combinations targeting urea, creatinine, and uric acid. The aim was to assess the biochemical effects, as well as the binding and sequestration efficiency, of these formulations under controlled experimental conditions. A randomized, double blind controlled in vitro study was conducted using serum samples obtained from twenty patients with uremia undergoing dialysis. Three formulations, labeled S1, S2, and S3, were evaluated.

All tested formulations resulted in statistically significant reductions in urea, creatinine, and uric acid concentrations compared with baseline values. Among them, the S1 formulation demonstrated the highest binding efficiency, reducing urea by 70% ± 7%, creatinine by 80% about 4%, and uric acid by 52% about 11%. Linear regression analysis confirmed a statistically significant association between the S1 formulation and reductions in these biochemical parameters.

These findings suggest that antibody functionalized granules can effectively bind and sequester nitrogenous waste products under in vitro conditions. This approach may represent a potential strategy for reducing uremic toxin burden, either as a complementary method or as a future alternative to existing renal replacement therapies. Further studies, including in vivo validation, dose optimization, and controlled clinical trials, are required to establish safety, efficacy, and translational applicability.

## Introduction

Chronic kidney disease (CKD) is a progressive and long-standing disorder characterized by a gradual decline in kidney function over time [1–4]. It is considered a major global health problem and one of the leading causes of morbidity and mortality worldwide [5–8]. The global burden of CKD is expected to increase further because of aging populations and the growing prevalence of major risk factors such as diabetes, hypertension, and obesity [9,10]. Although diabetes and hypertension are the most common causes of CKD globally, other factors such as glomerulonephritis, infections, environmental exposures, and genetic susceptibility also play important roles, particularly in developing countries [8,10–12].

CKD is generally defined by kidney damage or reduced kidney function persisting for more than 3 months, including decreased glomerular filtration rate (GFR), albuminuria, or other structural and functional abnormalities [13]. Early diagnosis and timely intervention are essential because they can delay disease progression, reduce complications, and improve quality of life [14–16]. Current management focuses on controlling the underlying cause, reducing cardiovascular risk, treating albuminuria, avoiding nephrotoxic agents, adjusting drug doses, and monitoring common complications such as anemia, metabolic acidosis, electrolyte imbalance, and mineral-bone disorders [13,17–19]. In patients at high risk of progression, referral to a nephrologist is strongly recommended [13].

Despite advances in supportive care, treatment options for advanced CKD remain limited. Dialysis and kidney transplantation are the standard therapies for end-stage disease, but both are expensive and may not be readily available in many settings [21,22]. Therefore, there is an ongoing need to develop alternative, supportive, and cost-effective therapeutic approaches that may reduce the toxic burden associated with renal failure.

One important concept in CKD pathophysiology is the close relationship between the kidneys and the intestine, commonly described as the gut–kidney axis [5,24]. This relationship is bidirectional and complex. The intestine contributes to the generation of several uremic toxins, especially those derived from protein metabolism and microbial activity [24,25]. In CKD and end-stage renal disease (ESRD), reduced renal excretion leads to the accumulation of toxic nitrogenous waste products in the blood, particularly urea, creatinine, and uric acid [14,26] as shown in **Supplementary Figure 1**.

Urea is the principal product of amino acid catabolism and is excreted mainly through the kidneys [27–29]. Creatinine is produced from muscle creatine metabolism and is widely used as a clinical marker of renal function because its blood level increases as GFR declines [28,31,32]. Uric acid, the product of purine metabolism, also accumulates in patients with impaired kidney function and may contribute to systemic complications [29,33]. Elevated concentrations of these nitrogenous metabolites are associated with fatigue, weakness, metabolic disturbances, and increased cardiovascular risk [14,34].

Recent studies have highlighted the important role of intestinal microbiota in CKD. The gut microbiome contributes to the production of several uremic retention solutes and their precursors, while microbial imbalance may promote inflammation and worsen toxin generation [35–40]. For this reason, several interventions such as dietary modification, probiotics, oral sorbents, and microbiome-targeted strategies have been investigated as possible methods to reduce toxin accumulation in CKD [41–44].

Although the kidneys and intestines have different primary physiological functions, both organs contribute to maintaining fluid balance, electrolyte homeostasis, and acid–base regulation [26,45,46]. This functional overlap has encouraged interest in the gastrointestinal tract as a possible auxiliary route for toxin removal in renal impairment. Experimental strategies such as oral sorbents, induced enteric excretion, and intestinal dialysis have been explored, although these approaches remain largely investigational and are not yet part of standard clinical practice [41,49–51,57,59,61].

The present study was designed to explore a novel enteric approach for reducing toxic nitrogenous waste accumulation in CKD. The proposed concept is based on capturing selected metabolites during their intestinal passage using targeted antibody-based binding systems, with the aim of lowering their systemic burden and reducing their harmful effects on renal tissue. On this basis, we developed PRO® A granules, a novel composite formulation intended to bind metabolic waste products within the gastrointestinal tract. As shown in **Supplementary figure 2 A, B**.

This formulation consists of four main components with complementary proposed actions. The first includes antibodies directed against urea, creatinine, and uric acid. he second contains goat antibody F(ab’)2 fragments against human IgA heavy chains, intended to enhance interaction with immune-related complexes associated with renal and mucosal pathology [25,63–67]. The third component is Pfu DNA polymerase proposed to support regenerative biological processes [68,69]. The fourth component is ispaghula husks a bulk-forming agent known to improve intestinal transit and potentially support enteric waste elimination [70] as shown in **Figure 2(A-D)**. In preliminary in vitro observations, 5 g of PRO® A granules showed the capacity to bind measurable amounts of creatinine, uric acid, and urea during simulated intestinal passage, suggesting a possible supportive role for this strategy in reducing nitrogenous toxin burden.

**Figure 1.**
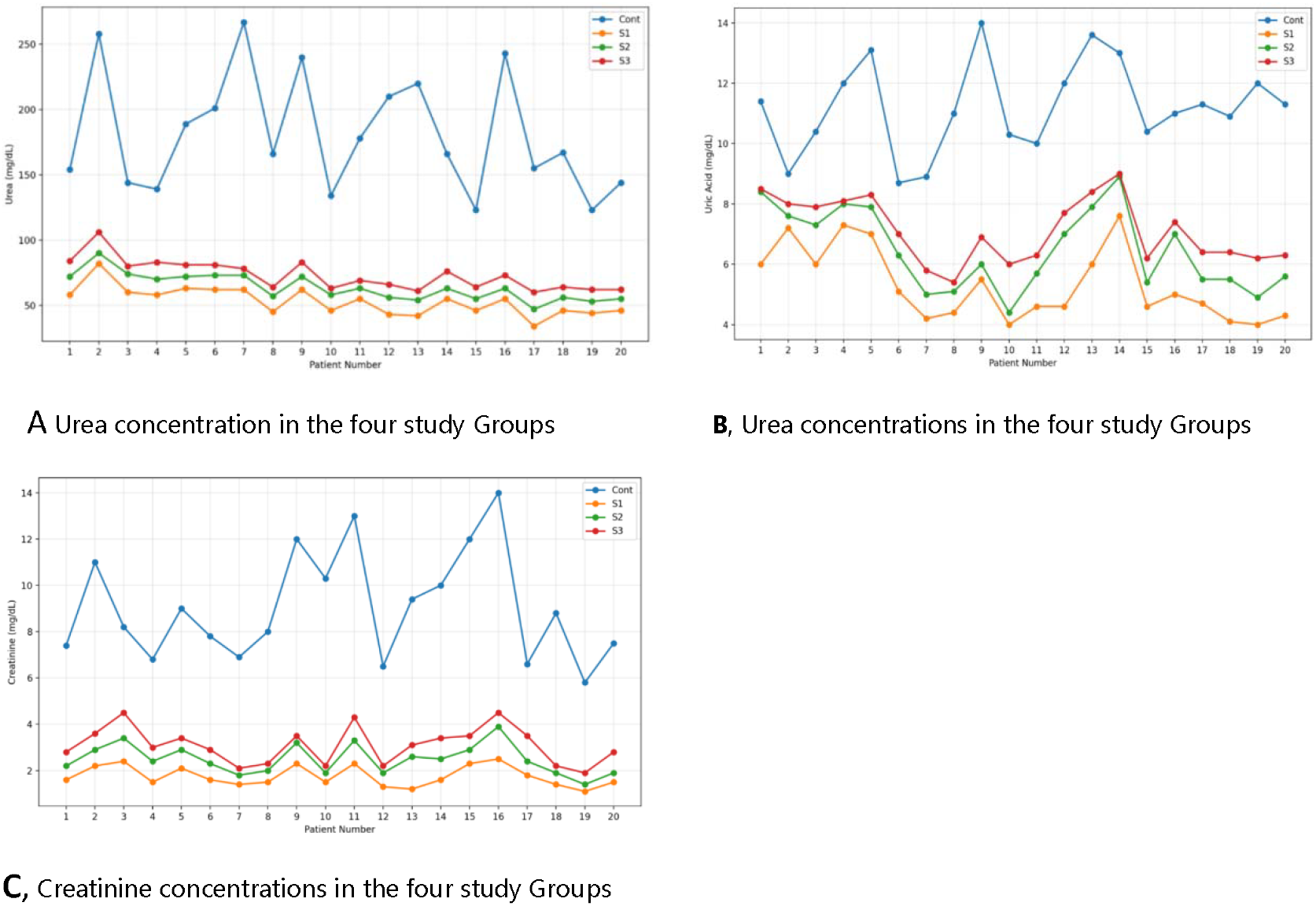
Distribution of nitrogenous waste product concentrations across study groups. (A) Urea concentrations, (B) creatinine concentrations, and (C) uric acid concentrations across individual patients under control conditions and following treatment with the tested formulations (S1–S3). All treated groups showed marked reductions compared with control, with Solution 1 demonstrating the greatest effect.

**Figure 2.**
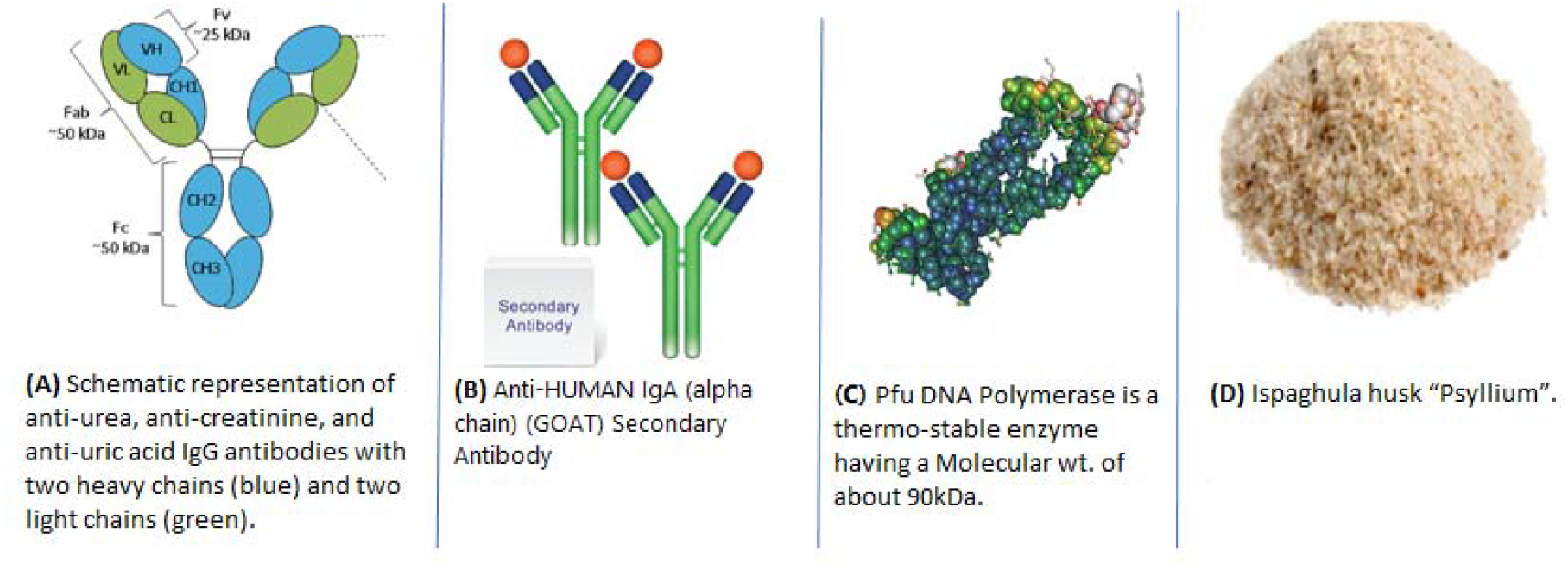
Functional components of PRO® A granules. (A–D) Schematic representation of the four principal components of the PRO® A granule formulation, including antibody-based targeting elements, enzymatic components, and the psyllium-based carrier matrix. These components are designed to support intestinal binding and sequestration of nitrogenous waste products.

## Materials and Methods

This study was designed as a pilot-scale, double-blind, controlled, in vitro preclinical investigation. Blood and urine samples were obtained from patients diagnosed with chronic kidney disease, with each participant serving as his or her own control. Untreated samples were used as baseline controls, whereas treated samples were exposed to the investigated granule formulation under standardized experimental conditions. The primary objective of this study was to evaluate a novel granule-based composition, PRO A granules, consisting of four functional components, for its capacity to bind and sequester nitrogenous waste products, including urea, creatinine, and uric acid, under simulated intestinal conditions. In addition,the study aimed to characterize the biochemical behavior of the tested formulation in biological samples obtained from patients with chronic kidney disease.

The study was conducted between 15 January 2025 and 20 February 2026 at the nephrology and hemodialysis outpatient unit of a private hospital in Cairo, Egypt. Samples were collected from twenty patients of both sexes diagnosed with chronic kidney disease at different stages, including patients with stage three and stage four disease, as well as patients undergoing maintenance hemodialysis. Eligible participants were between eighteen and seventy-five years of age and demonstrated significant biochemical impairment, defined by serum urea levels of at least one hundred fifty milligrams per deciliter, serum creatinine levels of at least seven milligrams per deciliter, and serum uric acid levels of at least twelve milligrams per deciliter. In addition, patients undergoing regular hemodialysis for a minimum duration of two years were considered eligible.

Participants were excluded if they were pregnant or breastfeeding, had received antibiotic therapy during screening or within the preceding fourteen days, declined to provide informed consent for the use of their samples in research, had active drug or alcohol dependence, were diagnosed with human immunodeficiency virus infection or significant liver disease, or were receiving ongoing anticoagulant therapy. All participants provided written informed consent prior to sample collection. Only samples obtained after confirmation of eligibility and completion of the consent process were included in the study.

### Materials

The investigated formulation was developed as a granule-based intestinal delivery system containing antibodies directed against selected uremic solutes, in combination with supportive components designed to enhance binding efficiency and delivery characteristics. The formulation included antibodies targeting urea, creatinine, and uric acid, each represented by immunoglobulin G monoclonal or polyclonal antibodies selected for their specificity and affinity toward their respective target molecules. Antibodies are widely used in biomedical applications because of their selective binding properties and high affinity for specific ligands [31–34]. In addition, a goat-derived antibody fragment, consisting of the antigen-binding fragment two directed against human immunoglobulin A, was incorporated as a functional component. Antibody fragments are commonly used in therapeutic and experimental systems because they retain antigen-binding capacity while providing advantages in terms of size, penetration, and functional flexibility [35–38].

The formulation also included a thermostable, high-fidelity deoxyribonucleic acid polymerase enzyme derived from Pyro coccus furious. This enzyme was incorporated as a ligand and potential enhancer of affinity between waste molecules and their corresponding antibodies, and as a component hypothesized to reduce interference from intestinal proteases [39–42].

Ispaghula husk, also known as psyllium, was used as the primary excipient and carrier matrix for the granule system. Psyllium is a natural dietary fiber characterized by its swelling and gel-forming properties and has been widely investigated for its use in pharmaceutical formulations and controlled delivery systems [43–49]. Stock preparations were generated from powdered antibody formulations and enzyme solutions. Antibodies directed against urea, creatinine, and uric acid, as well as goat immunoglobulin G directed against human immunoglobulin A, were prepared in defined concentrations, while the deoxyribonucleic acid polymerase enzyme was maintained in liquid form according to manufacturer specifications. Psyllium-based polymeric granules were synthesized and used as the carrier platform for incorporation of the active formulation. Previous studies have demonstrated that psyllium-based matrices can be modified for use in sustained-release and polymeric delivery systems [46–49].

Three experimental formulations were prepared by dissolving defined quantities of antibodies and supporting components in isotonic saline solution, followed by loading onto a fixed mass of granules. The relative concentrations of anti-urea, anti-creatinine, and anti-uric acid antibodies, together with the antibody fragment and enzyme component, were adjusted across the three formulations to evaluate concentration-dependent effects on binding efficiency.

The loading process was performed using a swelling equilibrium approach, in which the granules were exposed to the prepared solutions under controlled conditions, allowed to swell at physiological temperature, and subsequently dried to produce the final delivery system. Entrapment efficiency was estimated by quantifying the residual concentration of the loading solution after incorporation into the granule matrix [46,47].

In vitro evaluation was conducted using a simulated intestinal fluid prepared under standardized conditions and adjusted to a physiological pH. The medium was supplemented with defined concentrations of urea, creatinine, and uric acid to replicate uremic conditions. Granules loaded with each formulation were added to the simulated medium, while a control condition without granules was maintained for comparison. Samples were collected at multiple time points over a twenty-four-hour period to determine the kinetics of solute reduction and to identify the time required to reach equilibrium. The adsorption process was observed to stabilize at approximately six hours, and this time point was selected as the primary reference for subsequent analyses. Biochemical quantification of urea was performed using ultraviolet-visible spectrophotometry [50,51], creatinine was measured using the Jaffe colorimetric method [52], and uric acid levels were determined using column-switching liquid chromatography with ultraviolet detection [53].

Clinical sample collection included venous blood and urine specimens obtained from each participant. Blood samples were processed to obtain serum for biochemical analysis, including measurements of urea, creatinine, uric acid, alanine aminotransferase, and C-reactive protein. Urine samples were used for pregnancy testing when applicable and for assessment of protein and creatinine levels. To evaluate the functional efficiency of the antibody-loaded granules in biological matrices, serum samples derived from patients with chronic kidney disease were combined with simulated intestinal fluid and treated with the prepared formulations. Parallel untreated samples served as controls under standardized conditions. As shown in Figure 1A–C, concentrations of urea, creatinine, and uric acid were remeasured.

Statistical analysis was performed using appropriate parametric methods. Data were expressed as mean values with standard deviations, and comparisons between baseline and treated conditions were conducted using the paired-sample t test. Relationships between variables were further explored using linear regression analysis and curve estimation procedures implemented in the Statistical Package for the Social Sciences software, version twenty-five. A probability value of less than 0.05 was considered indicative of statistical significance.

## Results

A total of twenty patients diagnosed with chronic kidney disease were included in this pilot investigation. The age of the participants ranged from thirty-seven to seventy-five years, with a mean age of 53 ± 8.9 years. Body weight ranged from fifty-seven to one hundred seventeen kilograms, with a mean of 76 ± 18.7 kilograms, while height ranged from one hundred fifty-seven to one hundred seventy-six centimeters, with a mean value of 169.4 ± 6.3 centimeters. Male participants were more frequent than female participants, with fourteen males and six females included in the study. Most patients had a documented history of hypertension, and several individuals presented with additional comorbid conditions, including hyperlipidemia, type two diabetes mellitus, immunoglobulin A nephritis, polycystic kidney disease, focal segmental glomerulonephritis, mesangial proliferative glomerulonephritis, and systemic lupus erythematosus with renal involvement. All participants were receiving multiple pharmacological treatments for their underlying renal disease and associated systemic complications. As shown in Table 1.

**Table 1:**

| ID | Age (years) | Sex | Weight (kg) | Height (cm) | BMI (kg/m<sup>2</sup>) | Primary disease / relevant background condition |
| --- | --- | --- | --- | --- | --- | --- |
| 1 | 46–50 | F | 117 | 172 | 39.5 | CKD with associated hypertensive and metabolic conditions |
| 2 | 61–65 | M | 75 | 174 | 24.8 | CKD with associated hypertensive and metabolic conditions |
| 3 | 41–45 | M | 97 | 174 | 32.0 | CKD with associated hypertensive and metabolic conditions |
| 4 | 46–50 | F | 62 | 167 | 22.2 | CKD with associated hypertensive and metabolic conditions |
| 5 | 66–70 | M | 68 | 160 | 26.6 | CKD with associated hypertensive and metabolic conditions |
| 6 | 46–50 | F | 65 | 157 | 26.4 | CKD with associated hypertensive and metabolic conditions |
| 7 | 51–55 | M | 58 | 170 | 20.1 | CKD with associated hypertensive and metabolic conditions |
| 8 | 51–55 | M | 72 | 174 | 23.8 | CKD with associated hypertensive and metabolic conditions |
| 9 | 56–60 | M | 58 | 168 | 20.5 | CKD with associated hypertensive and metabolic conditions |
| 10 | 36–40 | M | 107 | 175 | 34.9 | CKD with associated hypertensive and metabolic conditions |
| 11 | 61–65 | M | 75 | 175 | 24.5 | CKD with associated hypertensive and metabolic conditions |
| 12 | 41–45 | M | 110 | 174 | 36.3 | CKD with associated hypertensive and metabolic conditions |
| 13 | 46–50 | F | 57 | 167 | 20.4 | CKD with associated hypertensive and metabolic conditions |
| 14 | 71–75 | M | 68 | 160 | 26.6 | CKD with associated hypertensive and metabolic conditions |
| 15 | 46–50 | F | 65 | 157 | 26.4 | CKD with associated hypertensive and metabolic conditions |
| 16 | 46–50 | M | 78 | 176 | 25.2 | CKD with associated hypertensive and metabolic conditions |
| 17 | 51–55 | M | 64 | 170 | 22.1 | CKD with associated hypertensive and metabolic conditions |
| 18 | 51–55 | M | 72 | 174 | 23.8 | CKD with associated hypertensive and metabolic conditions |
| 19 | 56–60 | F | 58 | 168 | 20.5 | CKD with associated hypertensive and metabolic conditions |
| 20 | 41–45 | M | 93 | 175 | 30.4 | CKD with associated hypertensive and metabolic conditions |

**Summary:** n = 20; Participants were distributed across multiple adult age categories; male/female: 14/6; BMI $26.4 \pm 5.6$ kg/m<sup>2</sup>.
**Abbreviations:** BMI, body mass index; F, female; M, male, CKD, chronic kidney disease.

Although a wide range of biochemical and clinical parameters were assessed, the present analysis focused primarily on serum urea, creatinine, and uric acid, as these represented the principal indicators used to evaluate the in vitro efficacy of the tested formulations. Three different formulations were investigated and subsequently loaded onto granules for testing under simulated intestinal conditions. The primary outcome measure was the reduction in the concentrations of urea, creatinine, and uric acid following exposure to the treated granules compared with untreated control samples.

The three tested formulations demonstrated clear binding and sequestration of nitrogenous waste products, whereas the control samples remained largely unchanged throughout the experimental period. The adsorption process was found to be time dependent, with equilibrium reached at approximately six hours, after which no meaningful additional reduction was observed at eight hours, indicating stabilization of the binding process.

Following eight hours of incubation, all treated samples showed substantial reductions in the concentrations of urea, creatinine, and uric acid compared with the control condition. The mean urea concentration after treatment was 53.20 ± 10.80 milligrams per deciliter with Solution 1, 63.80 ± 10.40 milligrams per deciliter with Solution 2, and 73.00 ± 11.66 milligrams per deciliter with Solution 3. In contrast, the mean urea concentration in the control group remained significantly higher at 181.05 ± 45.29 milligrams per deciliter.

Similarly, the mean creatinine concentration after treatment was reduced to 1.76 ± 0.44 milligrams per deciliter with Solution 1, 2.49 ± 0.65 milligrams per deciliter with Solution 2, and 3.09 ± 0.79 milligrams per deciliter with Solution 3, compared with a mean value of 9.05 ± 2.35 milligrams per deciliter in the control group. For uric acid, the mean concentration decreased to 5.31 ± 1.19 milligrams per deciliter with Solution 1, 6.47 ± 1.34 milligrams per deciliter with Solution 2, and 7.11 ± 1.06 milligrams per deciliter with Solution 3, whereas the control group maintained a mean concentration of 11.22 ± 1.50 milligrams per deciliter as shown in Figure 4A–C.

**Figure 3.**
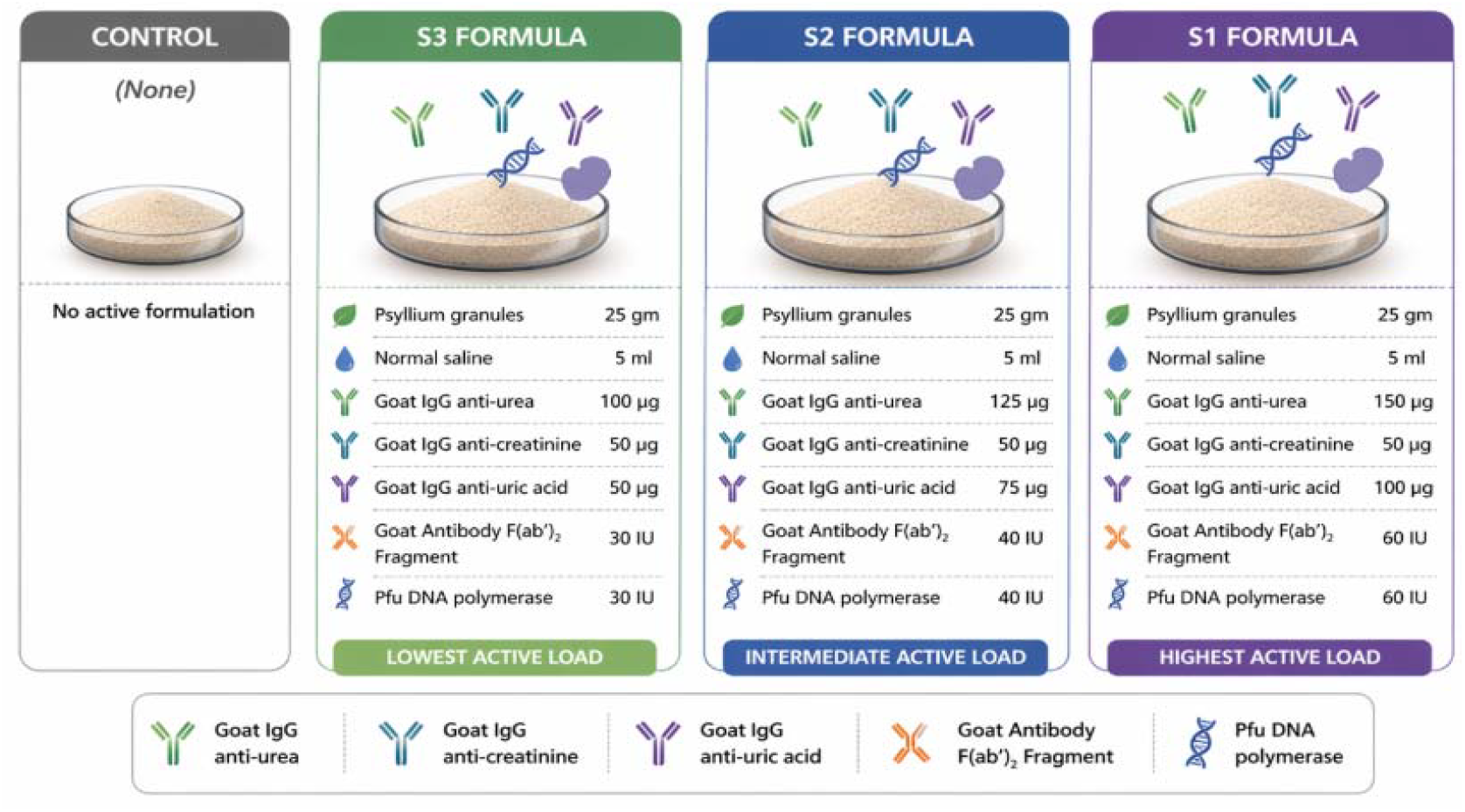
Schematic representation of the different formulations of PRO A granules used in the study. Each formulation consisted of psyllium-based granules loaded with varying concentrations of antibodies targeting urea, creatinine, and uric acid, in addition to supporting components. Solution 1 contained the highest concentrations, followed by Solution 2 and Solution 3, while the control group received no active formulation.

**Figure 4.**
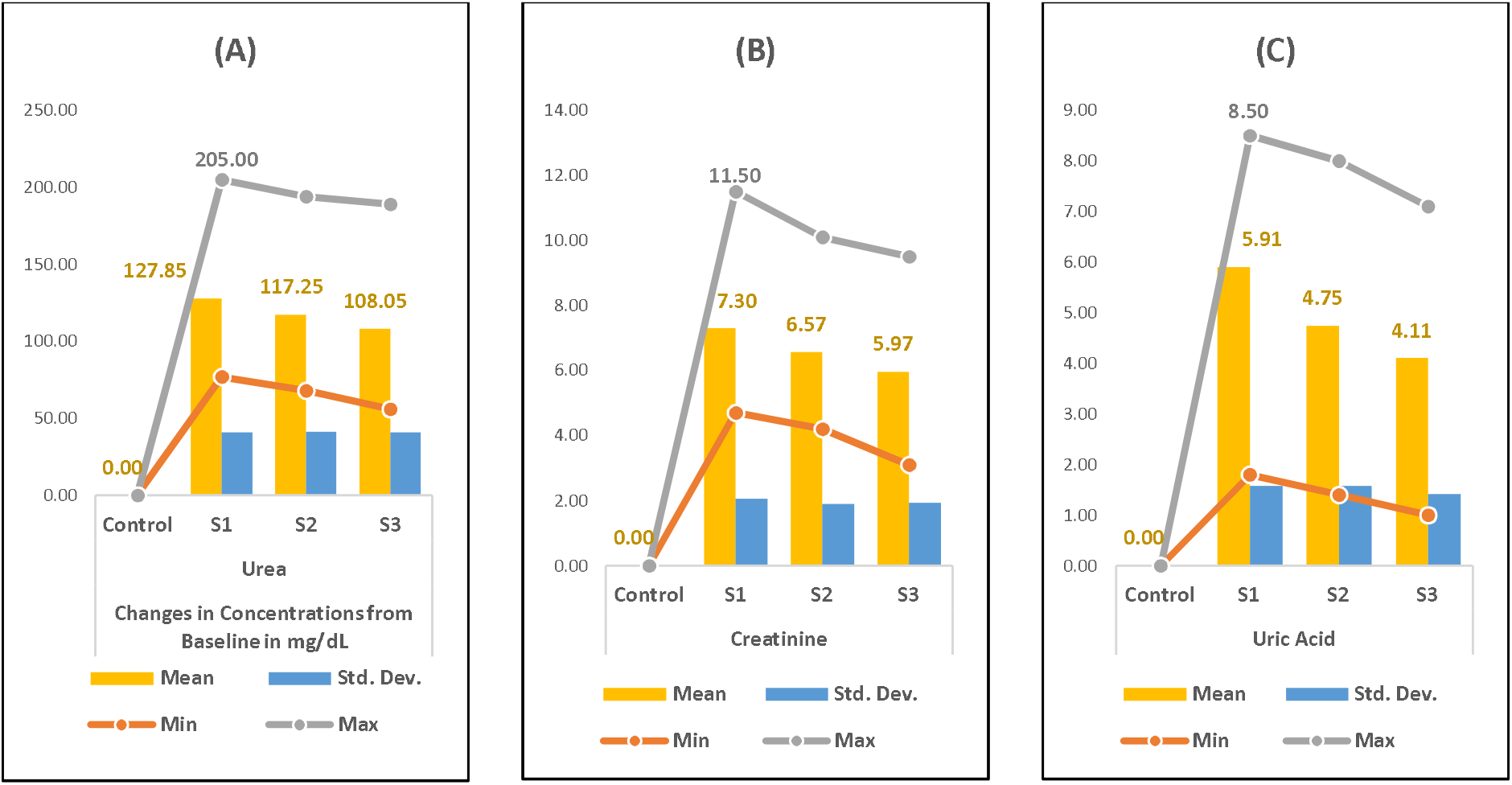
Statistical analysis of changes in nitrogenous waste product concentrations. (A) Urea, (B) creatinine, and (C) uric acid levels following treatment with the tested formulations. Data are expressed as mean ± standard deviation. All treatment groups demonstrated statistically significant reductions compared with control.

When the reduction from baseline values was examined, no significant change was observed in the control samples, whereas all treated formulations produced measurable reductions in the studied analytes. Solution 1 demonstrated the strongest overall effect, with urea reduction ranging from seventy-seven to two hundred five milligrams per deciliter and a mean reduction of 127.85 ± 40.79 milligrams per deciliter, corresponding to an average reduction of 70% ± 7%. Creatinine reduction ranged from 4.7 to 11.5 milligrams per deciliter, with a mean reduction of 7.3 ± 2.05 milligrams per deciliter, corresponding to an average reduction of 80% ± 4%. Uric acid reduction ranged from 1.8 to 8.5 milligrams per deciliter, with a mean reduction of 5.91 ± 1.58 milligrams per deciliter, corresponding to an average reduction of 52% ± 11%. Solution 2 showed intermediate activity, with a mean urea reduction of 117.25 ± 41.16 milligrams per deciliter, corresponding to approximately 63% ± 8%, a mean creatinine reduction of 6.57 ± 1.91 milligrams per deciliter, corresponding to approximately 72% ± 5%, and a mean uric acid reduction of 4.75 ± 1.58 milligrams per deciliter, corresponding to approximately 42% ± 12%.

Solution 3 demonstrated the lowest, yet still evident, activity, with a mean urea reduction of 108.05 ± 41.09 milligrams per deciliter, corresponding to approximately 58% ± 9%, a mean creatinine reduction of 5.97 ± 1.94 milligrams per deciliter, corresponding to approximately 65% ± 8%, and a mean uric acid reduction of 4.11 ± 1.42 milligrams per deciliter, corresponding to approximately 36% ± 10%. Among the three formulations, Solution 1 consistently achieved the greatest reduction in all measured analytes. The maximum observed reduction in creatinine reached 11.5 milligrams per deciliter, corresponding to an eighty-seven percent decrease in certain samples, while urea reduction reached up to 205 milligrams per deciliter, corresponding to an eighty-one percent decrease.

All three formulations produced statistically significant reductions compared with baseline values based on paired-sample t test analysis at a ninety-five percent confidence level. Because Solution 1 demonstrated superior performance, it was selected for further regression analysis. For urea, the linear relationship between baseline and post-treatment values was statistically significant, with the equation y = 0.1229x + 30.945, an F value of 6.51, a probability value of 0.02, and a coefficient of determination of 0.2656. For creatinine, the linear relationship was also statistically significant and demonstrated a stronger model fit, with the equation y = 0.137x + 0.5153, an F value of 20.49, a probability value of 0.00, and a coefficient of determination of 0.5323. In contrast, the regression model for uric acid showed greater variability and did not reach statistical significance, with the equation y = 0.2599x + 2.3953, an F value of 2.133, a probability value of 0.16, and a coefficient of determination of 0.106. As showed in **Figure 5**.

**Figure 5.**
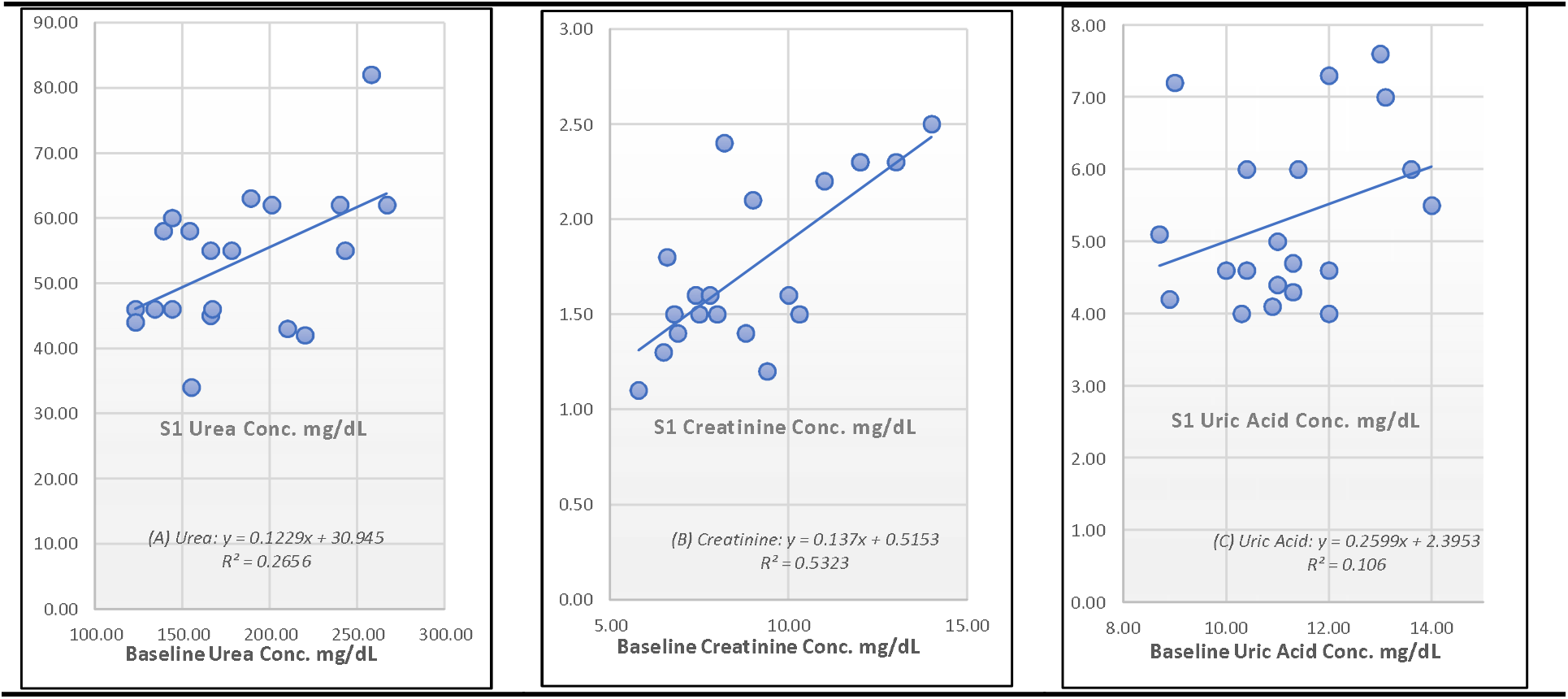
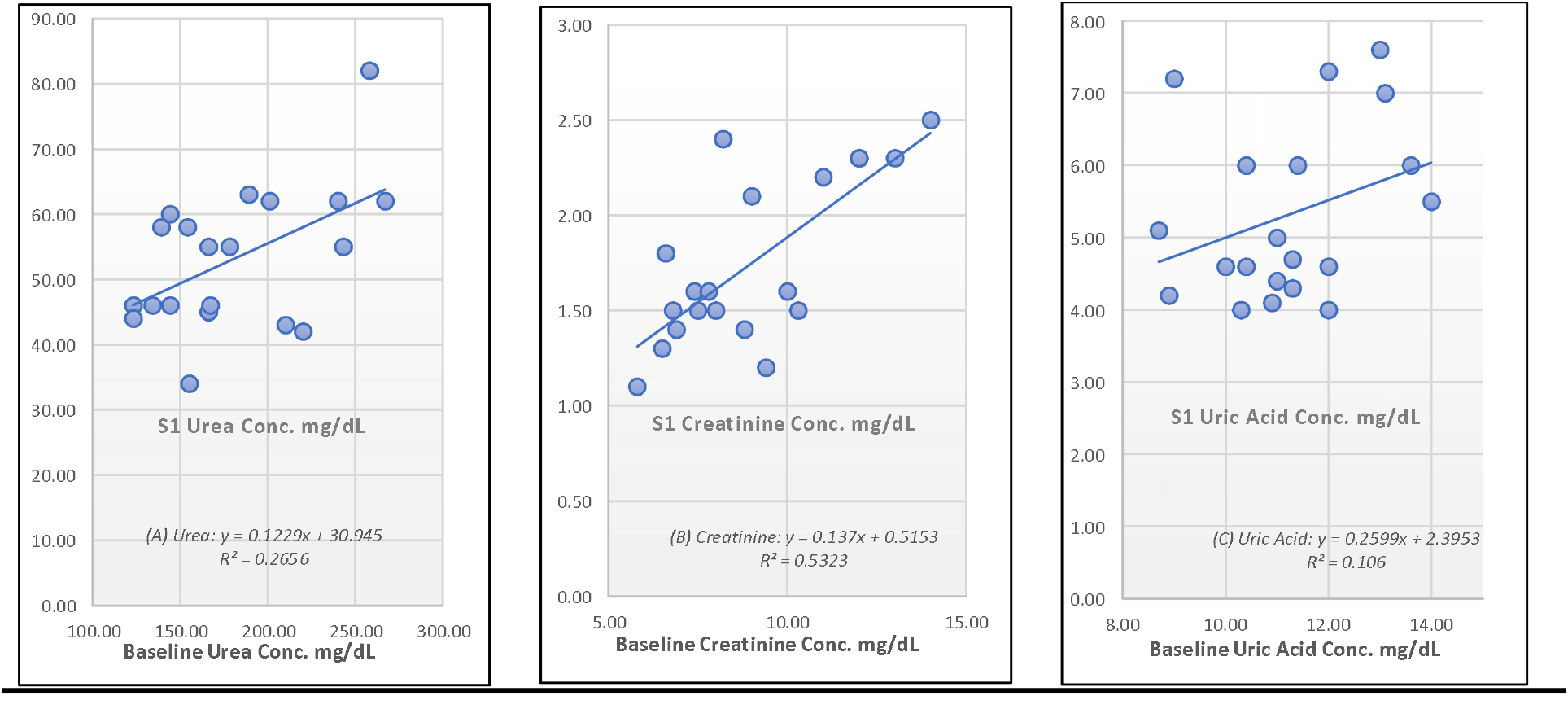
Normal probability–probability (P–P) plot of regression standardized residuals for nitrogenous waste reduction using Solution 1. The plot demonstrates the normal distribution of residuals, supporting the validity of the regression model applied to evaluate the relationship between baseline and post-treatment values.

Overall, the findings demonstrated that the tested granule formulation could reduce the concentrations of urea, creatinine, and uric acid under simulated intestinal conditions. The observed effect was dependent on the composition and concentration of the formulation, with Solution 1 showing the highest efficacy across all evaluated parameters. These results support further investigation in experimental animal models and future clinical evaluation.

## Discussion

chronic kidney disease (CKD) remains a major clinical and public health challenge because of its progressive nature, its close association with metabolic complications, and the high cost of advanced treatment options. In many patients, progressive loss of renal function leads to retention of nitrogenous waste products such as urea, creatinine, and uric acid, which contribute directly to systemic complications and worsening quality of life [31–33]. Although dialysis and kidney transplantation remain the main therapeutic approaches for advanced disease, these options are not always easily accessible and may be associated with significant physical, economic, and logistical burdens [34]. The current study was designed to explore an alternative supportive strategy based on the concept of enteric waste reduction. This concept is supported by the physiological role of the intestine as a large semipermeable surface that can participate in the movement and handling of small solutes under certain pathological conditions [35,36]. In uremic states, the gastrointestinal tract may partially contribute to the clearance of retained metabolites, which provides a scientific rationale for the development of intestinal binding systems aimed at reducing circulating waste products [37].

In the present work, the tested formulation demonstrated a measurable in vitro capacity to bind and reduce key nitrogenous waste products. Among the evaluated preparations, formula S1 showed the most favorable performance, with the highest reduction percentages for urea, creatinine, and uric acid. This finding suggests that the selected composition may provide more efficient adsorption characteristics than the other tested formulas. In addition, the observed stabilization of binding after approximately 6 hours indicates that the interaction between the granules and the targeted solutes reaches a relatively consistent equilibrium under the study conditions. An important observation in this study is the significant linear relationship between baseline concentrations of the measured waste products and the extent of reduction achieved by the formula. This may indicate that the adsorption system becomes more effective in samples with a higher waste burden, which could be particularly relevant in patients with more advanced renal dysfunction. From a therapeutic perspective, this result supports the possibility that such an intestinal approach may have practical value in reducing the biochemical burden associated with CKD, especially when conventional renal replacement therapy is delayed or limited. The significance of these findings lies in the fact that current conservative management of CKD is mainly directed toward slowing disease progression and controlling complications rather than directly reducing intestinally available nitrogenous waste. Therefore, a formulation capable of acting locally in the gut and binding uremic solutes may represent a useful adjunctive strategy rather than a replacement for standard therapy. If future in vivo studies confirm these results, this approach may help reduce metabolic stress and improve biochemical control in selected patients [38].

The formulation strategy itself may also represent an advantage. The use of psyllium-based granules provides a practical carrier with known medical use and generally acceptable tolerability. At the same time, the inclusion of specific binding components appears to enhance the ability of the system to adsorb target waste products. However, despite these promising preliminary results, safety remains an essential concern. The gastrointestinal behavior of the incorporated antibodies and related components, as well as their stability, tolerability, and possible biological interactions after oral administration, require further careful study before translation into clinical application can be justified [39].

This study has several limitations that should be acknowledged. First, it is an in vitro pilot study, and therefore the experimental setting does not completely reproduce the complex intestinal environment found in vivo. Factors such as intestinal motility, pH variability, digestive processes, microbiota, mucus interaction, and transit time may influence performance under real biological conditions. Second, the sample size was limited, which means that the results should be interpreted as preliminary evidence rather than definitive proof of efficacy. Third, although the biochemical reductions were encouraging, the study does not establish whether these effects would translate into meaningful clinical outcomes such as delaying dialysis, improving symptoms, or reducing systemic complications. Despite these limitations, the findings provide a useful basis for future research. The most immediate next step would be to evaluate the formulation in animal models and then in controlled human studies to determine its safety, tolerability, and real therapeutic value. Further optimization of the formulation may also improve adsorption efficiency and consistency. In addition, this approach may have broader implications in disorders characterized by metabolite retention or altered intestinal solute handling, although such applications remain speculative at present [40].

In conclusion, the present study provides preliminary support for the concept that PRO® A granules, particularly formula S1, can effectively bind major nitrogenous waste products in vitro. These findings strengthen the idea that enteric waste capture may represent a promising non-invasive adjunctive strategy in CKD management. However, substantial further work is still needed to confirm safety, biological feasibility, and clinical relevance before this approach can be considered for practical therapeutic use.

## Conclusion

In conclusion, our study demonstrates that the novel formulas of antibodies combinations that we have invented can effectively bind and sequestrate nitrogenous waste products *in vitro*. The results of our experiments indicate that the (S1) formula has the highest binding efficiency, reducing urea by 70%±7%, creatinine by 80%±4%, and uric acid by 52%±11%. The linear regression analysis suggests that the reductions in nitrogenous waste products are linearly related to the baseline concentrations of these substances in the patient’s serum. We suggest further research in animal and clinical trials to confirm the accuracy, efficiency, and safety of these formulas for the treatment of uremic patients.

## Data Availability

All data produced in the present work are contained in the manuscript

https://doi.org/10.5281/zenodo.19747217

## Institutional Review Board Statement

The study protocol was reviewed and approved by the Institutional Review Board of the participating clinical facility, in accordance with national and institutional ethical guidelines.

## Ethical Approval Statement

The study protocol was reviewed and approved by the Institutional Review Board (IRB) of Al-Qasr Al-Aini Hospital, Cairo, Egypt. The study was conducted in accordance with the ethical standards of the Declaration of Helsinki and applicable national regulations.

## Informed Consent Statement

All authors have read and agreed to the published version of the manuscript.

## Data Availability Statement

The data that support the findings of this study are available on request from the corresponding authors.

## Author Contributions

Authors’ Contributions are not equal. S. Salah conceived the study; S. Sherif, M. Abdula wrote the manuscript & designed the experiments. M Abdula performed the experiments, analyzed the data.

## Supplementary Materials

Additional datasets supporting this study are available at:

https://doi.org/10.5281/zenodo.19747217

